# Psychosocial Risk Assessment Among Mental Health Workers in a Regional Psychiatric Hospital

**DOI:** 10.1101/2025.08.12.25333502

**Authors:** Elodie Canut, Yvonne Donati, Chloé Hiver, Dominique Pauvarel, Antoine Villa, Marie-Pascale Lehucher-Michel

## Abstract

This study aimed to assess ‘job strain” (JS) and effort-reward imbalance (ERI), two markers of work-related stress, among psychiatric healthcare workers exposed to patients’ psychological suffering. A self-administered questionnaire was distributed in a regional French public hospital. The response rate was 65% and the prevalence of JS was estimated at 37%. Occurrence of JS was increased in the inpatient sector (OR 1.94 [1.01-3.74], P(χ²)=0.046), decreased by day work (OR 0.53 [0.29-0.95], P(χ²)=0.032), and by larger hospital seniority. The ERI ratio (mean 0.537 ± 0.228) was higher among psychiatrists and workers doing overtime. It was lower in staff who felt supported during episodes of violence. These results suggest the need for specific preventive measures targeting these risk factors.

## Introduction

During the past two decades, psychosocial risks (PSRs) have steadily increased and have become a major public health issue in France (1) and worldwide (2). The cost of work-related stress in France was estimated between 2 and 3 billion euros in 2007 (3). The “job strain” (4) and the effort-reward imbalance (5) are markers of stress at work and have been linked to an increased risk of cardiovascular disease (6,7), mental troubles (8) and musculoskeletal disorders (9). PSRs also result in higher rates of sick leave, absenteeism, and increased turnover (1).

Healthcare workers have a high risk of stress worldwide (10–13). Physicians and nurses are particularly prone to work rate constraints, task interruptions, and emotional demands due to the nature of their mission. Over the past decade, they report more frequent assaults from the public (14). Stress-related staff shortage due to absenteeism or sickness absence reports the workload over the remaining workers, overburdening the healthcare staff. Such difficulties may hamper job attractivity (15).

In psychiatric units, healthcare workers are exposed to patients’ psychological more than physical suffering, which could increase the occurrence of PSRs. However the staff is trained to care for mental illness and psychological disorders and may thus develop protective skills. For instance, a German study reported higher levels of burnout and a greater effort-reward imbalance among nurses working in somatic compared with psychiatric hospital settings (16).

In France, previous research has addressed the burnout of healthcare workers (10,17) and mental health professionals in psychiatry (18). A qualitative study on the quality of life at work for psychiatrists in hospitals has also been carried out (19). However, to our knowledge, “job strain” has never been quantitatively investigated in psychiatric healthcare workers in hospitals.

The aim of this study was to assess the prevalence of “job strain” in this population and analyze the factors that may influence its occurrence.

## Materials and Methods

### Target population

The target population consisted of 215 healthcare workers (176 nurses, 30 physicians, and 9 paramedical supervisors) composing an adult psychiatry department in a regional public hospital of southern France. The distribution of healthcare workers within the adult psychiatry department is presented in Table 1.

**Table 1:** Distribution of healthcare staff in the adult psychiatry department.

| Unit of the department | Staff (workers) |
| --- | --- |
| *Psychiatry in full-time hospitalization (162 beds)* |  |
| 5 open hospitalization units | 91 |
| 1 addiction treatment unit | 13 |
| 3 closed hospitalization units and an emergency service | 56 |
| *Ambulatory psychiatric structures* |  |
| 4 medico-psychological centers | 26 |
| 2 day hospitals and 2 part-time therapeutic reception centers | 18 |
| Therapeutic-oriented associative apartments | 6 |
| 2 mobile psychiatry teams for precariousness | 5 |

From October 8^th^, 2023, to January 12^th^, 2024, a self-administered questionnaire was introduced and distributed to the target population by an occupational physician. Two to five follow-up reminders were made within the units.

### Content of the self-administered questionnaire

The self-administered questionnaire consisted of:

i. A custom-made questionnaire with various items regarding: profession type, seniority, employment contract settings, anthropometric data, parenthood, health status, history of work-related accidents or occupational diseases, tobacco, alcohol and psychoactive substance use, as well as information on specific medical and paramedical training in psychiatry. The synthesis meeting is a multidisciplinary meeting of professionals involved in patients’ care, allowing for discussion and coordination in support of the therapeutic project. The supervision meeting is organized by a professional not directly involved in patients’ care, providing a time for listening and expressing regarding the experiences of healthcare workers in their practice.
ii. The Karasek questionnaire, assessing “job strain” through the combination of high psychological demand and low decision latitude. We used decision latitude threshold scores of 70.3 and psychological demand of 21, as defined in the French employee population (Sumer survey (20)). This questionnaire also evaluates social support through an independent score.
iii. The Siegrist questionnaire, measuring the effort-reward imbalance at work through a numerical ratio. This questionnaire also evaluates work overinvestment through an independent score.

### Ethics

This workplace action has been conducted in the target population as part of routine occupational health surveillance. Resulting anonymized data were collected after local ethics committee approval (IRB #00012962). The data were analyzed in accordance with the Helsinki Declaration.

### Treatment of missing data

Missing data from the Karasek and Siegrist questionnaires were filled in according to the following completion procedure: for an unanswered question of weight W, the theoretical median value was assigned if W was smaller than the standard deviation (σ) of the group in that category. Thus, the goal was to avoid substantial alteration of the questionnaire result, with respect to the distribution of values within the rest of the sample. For example, if data was missing in the decision latitude category for a question weighing 4 to 16 points, a value of 10 was assigned if 16 was smaller than the group’s standard deviation in that category. If two or more questions were unanswered in the same category, their weights added up and may rapidly exceed σ. If W > σ, the entire category was neutralized and kept as missing data.

### Statistical analysis

Qualitative variables are expressed as the absolute number of workers and as the percentage over the total number of responses collected. Quantitative variables are expressed as the mean ± standard deviation in the sample.

Statistical analysis was performed using Excel® (Microsoft). The significance of the relationship between the collected variables and “job strain” occurrence was assessed using a χ² test for qualitative variables. The strength of the relationship was determined by the odds ratio (OR) and the confidence interval calculated from the χ² statistic. For quantitative variables, groups were compared using a bilateral paired Student’s t-test with Welch’s correction in case of unequal variances. For analyzing the relationship between quantitative variables and the quantitative ratio of effort-reward imbalance, the Pearson correlation coefficient was used. A p-value or χ² < 0.05 was considered statistically significant.

## Results

In this study, 140 out of 215 targeted staff members filled in the questionnaire, resulting in a participation rate of 65.1%.

Before the completion procedure, 19 Karasek and Siegrist questionnaires had missing data. After completion, the Karasek score was assessed in 139 participants (99.3%), and the effort-reward imbalance ratio in 134 participants (95.7%).

Table 2 presents descriptive data of the studied population.

**Table 2:** Descriptive data of the studied population.

| **Variables** | **N (%) or mean**<br>**± SD** |
| --- | --- |
| Female | 91 (65.0) |
| Age (years) | 45.8 ± 11.2 |
| Seniority (years) | 14.5 ± 10.6 |
| Parenthood | 104 (75.4) |
| Body-mass index (kg/m<sup>2</sup>) | 24.0 ± 4.1 |
| Hypertension | 6 (4.4) |
| Diabetes | 2 (1.5) |
| Hypercholesterolemia | 6 (4.4) |
| Work-related accident or occupational disease | 45 (32.1) |
| History of burnout | 28 (21.2) |
| History of psychiatric disorders | 32 (23.2) |
| Requiring treatment | 30 (22.9) |
| Self-reported good health | 123 (87.9) |
| Smoking status | 30 (21.7) |
| Alcohol | 94 (67.1) |
| <u>*Alcohol consumption*</u> |  |
| 0 | 46 (32.9) |
| Rarely | 25 (17.9) |
| Once/month | 18 (12.9) |
| Once/week | 49 (35.0) |
| Once/day | 2 (1.4) |
| Coffee or tea | 110 (78.6) |
| <u>*Coffee or tea consumption*</u> |  |
| 0 | 30 (21.6) |
| Once/day | 21 (15.1) |
| Twice/day | 33 (23.7) |
| 3 to 5 times/day | 45 (32.4) |
| More than 5 times/day | 10 (7.2) |
| Energy drinks | 4 (2.9) |
| *<u>Profession</u>* |  |
| Psychiatrist | 15 (10.7) |
| Paramedical supervisor | 7 (5.0) |
| Nurses | 112 (80.0) |
| Non-psychiatrist physician | 6 (4.3) |
| Work ability restriction | 10 (7.4) |
| Permanent staff | 127 (90.7) |
| <u>Work schedule</u> |  |
| 100% | 119 (85.6) |
| 90% | 2 (1.4) |
| 80% | 17 (12.2) |
| 60% | 1 (0.7) |
| Overtime | 85 (62.0) |
| <u>Number of overtime hours</u> |  |
| 0 | 53 (39.0) |
| 0 to 5 hours/month | 39 (28.7) |
| 5 to 10 hours/month | 26 (19.1) |
| More than 10 hours/month | 18 (13.2) |
| <u>Hospital sector</u> |  |
| Inpatient | 92 (65.7) |
| Outpatient | 32 (22.9) |
| Mixed | 16 (11.4) |
| <u>Work period</u> |  |
| Day work | 61 (43.6) |
| Nigth work | 10 (7.1) |
| Mixed | 69 (49.3) |
| Choice of psychiatry | 137 (97.9) |
| Supervisors | 131 (93.6) |
| Ethical questioning | 101 (80.1) |
| Training in care | 92 (66.2) |
| Training in violence management | 69 (49.3) |
| Feeling supported during episodes of violence | 93 (68.4) |
| Training in care without consent | 108 (78.3) |
| Synthesis meetings | 104 (75.9) |
| Supervision meetings | 71 (51.5) |
| *<u>Karasek Categories</u>* |  |
| Low strain | 13 (9.4) |
| Active work | 65 (46.8) |
| Passive work | 9 (6.5) |
| **High strain (« job strain »)** | **52 (37.4)** |
| Siegrist effort-reward imbalance ratio | 0.537 ± 0.228 |
| Siegrist ratio above 1 | 6 (4.5) |

Within respondents, 65% were women, with an average age of 45.8 ± 11.2 years and an average seniority of 14.5 ± 10.6 years. 75.4% had children.

In the population, 80% were nurses, 85.6% worked full-time, and 62% reported doing overtime (71.4% in the inpatient sector, 43.5% in the outpatient or mixed sector).

The staff were distributed as follows: 65.7% in the inpatient sector, 22.9% exclusively in the outpatient sector, and 11.4% in a mixed sector.

A portion of 43.6% worked only during the day, 7.1% worked only at night, and 49.3% had a mixed day-night work schedule.

87.9% of respondents considered themselves in good health, 32.1% had experienced a work-related accident or occupational disease, and 21.2% had experienced burnout.

The smoking rate was 21.7%, while 67.1% consumed alcohol, and 78.6% drank coffee or tea, with 39.6% consuming it more than three times per day.

23.2% of respondents had a psychiatric disorder history, with 22.9% of them receiving treatment.

Among respondents, 49.3% reported receiving training in managing violence. 75.9% participated in synthesis meetings, and 51.5% participated in supervision meetings.

The prevalence of “job strain” in the studied population was 37.4% [29.4 – 45.4%]. The comparison of individual Karasek questionnaire scores in the population with the thresholds from the Sumer survey is shown in Figure 1.

**Figure 1:**
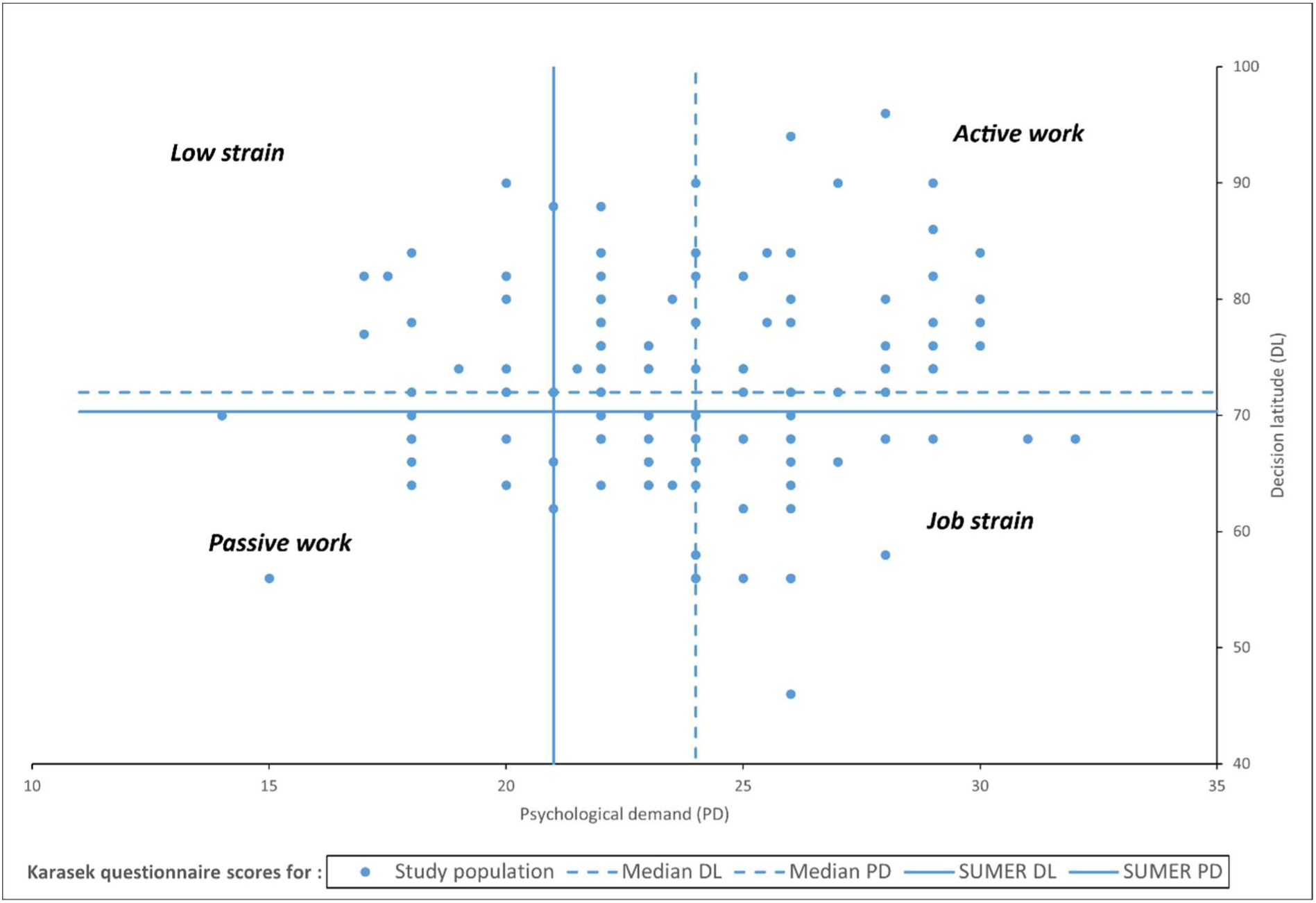
Karasek questionnaire scores plotted for each respondent. Median values in the population are represented as dashed lines. Threshold values from the Sumer 2003 survey are represented as solid lines.

The effort-reward imbalance ratio had an average value of 0.537 ± 0.228, and was greater than 1 for 6 respondents (4.5%).

The association of “job strain” with the characteristics of the studied population in univariate analysis is detailed in Table 3. The 16 most relevant variables for analyzing “job strain” are summarized in Figure 2: overtime, inpatient work, day work, female gender, parenthood, self-reported good health, history of work-related accidents or occupational diseases, history of burnout, history of psychiatric disorder, smoking, alcohol consumption > 1/week, coffee or tea consumption > 3/day, training in violence de-escalation, help during violence episodes, synthesis meetings, and supervision meetings.

**Figure 2:**
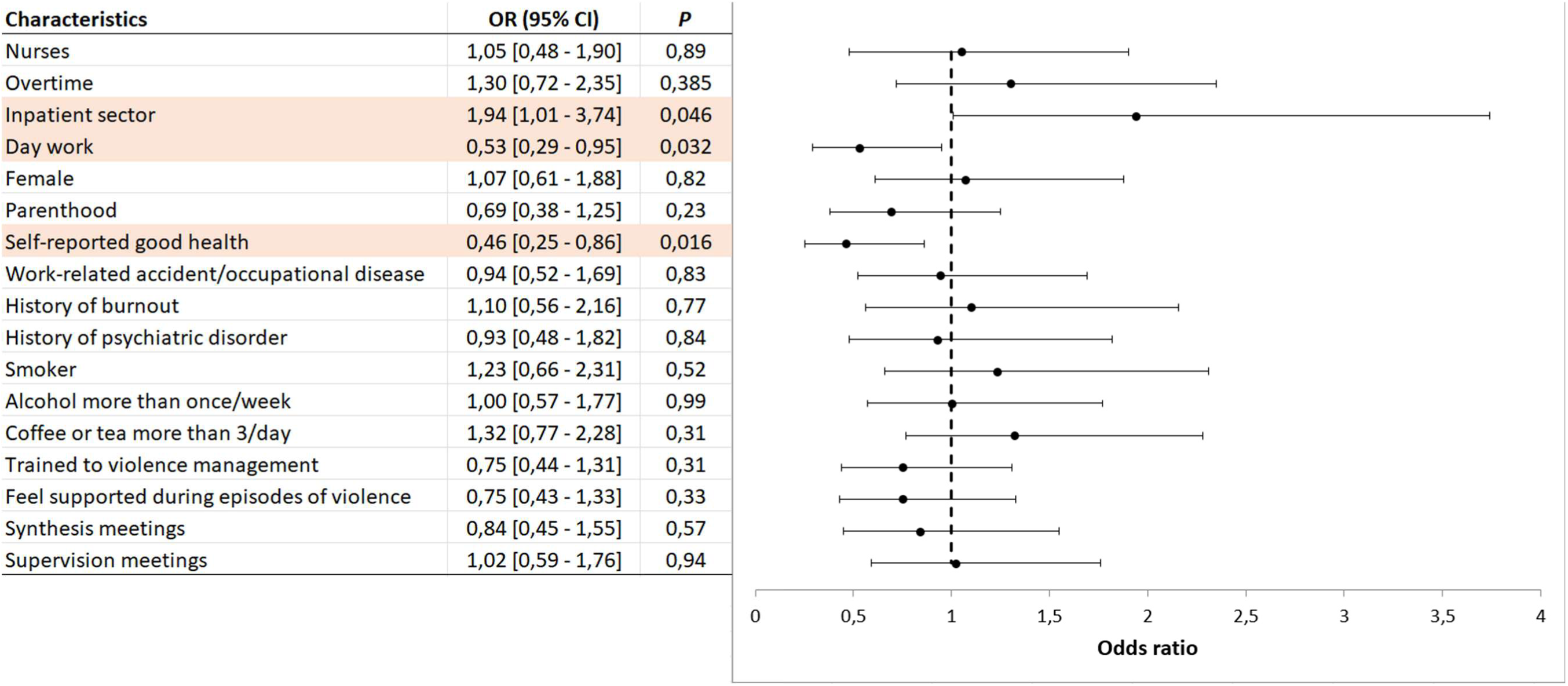
Odds ratio for the occurrence of « job strain » for 16 variables in the studied population.

**Table 3:** association of “job strain” with the characteristics of the studied population in univariate analysis.

| **Characteristics** | **Job strain**<br>N (%) or<br>mean<br>± SD | **Other**<br>**categories**<br>N (%) or<br>mean<br>± SD | **OR (95% CI)** | **P ($\chi^2$)** |
| --- | --- | --- | --- | --- |
| Healthcare workers | 52 (37.4) | 87 (62.6) |  |  |
| Psychiatrist | 6 (40.0) | 46 (36.8) | 1.09 [0.40 - 2.97] | 0.85 |
| Paramedical supervisor | 2 (28.6) | 50 (37.6) | 0.76 [0.15 - 3.78] | 0.7 |
| Nurse | 42 (37.5) | 10 (35.7) | 1.05 [0.47 - 2.35] | 0.89 |
| Non-psychiatrist physician | 2 (33.3) | 50 (37.3) | 0.89 [0.17 - 4.57] | 0.88 |
| Overtime | 34 (40.0) | 16 (30.8) | 1.30 [0.65 - 2.58] | 0.385 |
| Inpatient sector | 41 (44.6) | 11 (22.9) | 1.94 [0.92 - 4.12] | 0.046 |
| Day work | 15 (24.6) | 37 (46.8) | 0.53 [0.26 - 1.04] | 0.032 |
| Seniority (years) | 12.1 ± 9.5 | 16.0 ± 11.0 |  | 0.03 |
| Age (years) | 43.6 ± 10.7 | 47.0 ± 11.3 |  | 0.08 |
| Body-mass index (kg/m<sup>2</sup>) | 24.2 ± 4.0 | 23.8 ± 4.2 |  | 0.58 |
| Female | 33 (36.3) | 19 (38.8) | 1.07 [0.55 - 2.07] | 0.82 |
| Parenthood | 34 (32.7) | 16 (47.1) | 0.69 [0.34 - 1.41] | 0.23 |
| Self-reported good health | 40 (32.5) | 12 (70.6) | 0.46 [0.20 - 1.05] | 0.016 |
| Work-related accident/occupational disease | 16 (35.6) | 36 (37.9) | 0.94 [0.47 - 1.87] | 0.83 |
| History of burnout | 11 (39.3) | 37 (35.6) | 1.10 [0.50 - 2.44] | 0.77 |
| Smoker | 13 (43.3) | 38 (35.2) | 1.23 [0.58 - 2.60] | 0.52 |
| Alcohol | 33 (35.1) | 19 (41.3) | 0.85 [0.44 - 1.65] | 0.57 |
| Alcohol more than 1/week | 19 (37.3) | 33 (37.1) | 1.00 [0.52 - 1.95] | 0.99 |
| Coffe or tea | 40 (36.4) | 12 (40.0) | 0.91 [0.42 - 1.95] | 0.77 |
| Coffee or tea more than 3 times/day | 24 (43.6) | 28 (32.9) | 1.32 [0.70 - 2.52] | 0.31 |
| History of psychiatric disorders | 11 (34.4) | 39 (36.8) | 0.93 [0.43 - 2.03] | 0.84 |
| Training in violence management | 22 (31.9) | 30 (42.3) | 0.75 [0.40 - 1.43] | 0.31 |
| Feeling supported during episodes of violence | 31 (33.3) | 19 (44.2) | 0.75 [0.38 - 1.48] | 0.33 |
| Synthesis meetings | 37 (35.6) | 14 (42.4) | 0.84 [0.40 - 1.74] | 0.57 |
| Supervision meetings | 27 (38.0) | 25 (37.3) | 1.02 [0.54 - 1.93] | 0.94 |

Working in the inpatient sector was identified as a risk factor (OR 1.94 [1.01-3.74], P(χ²)=0.046).

Working only during the day (OR 0.53 [0.29-0.95], P(χ²)=0.032), longer average seniority (12.1 ± 9.5 years *vs* 16.0 ± 11.1 years, P=0.03), and self-reported good health (OR 0.46 [0.25-0.86], P(χ²)=0.016) were protective factors.

The effort-reward imbalance ratio in population characteristics subgroups is presented in Table 4.

**Table 4:**
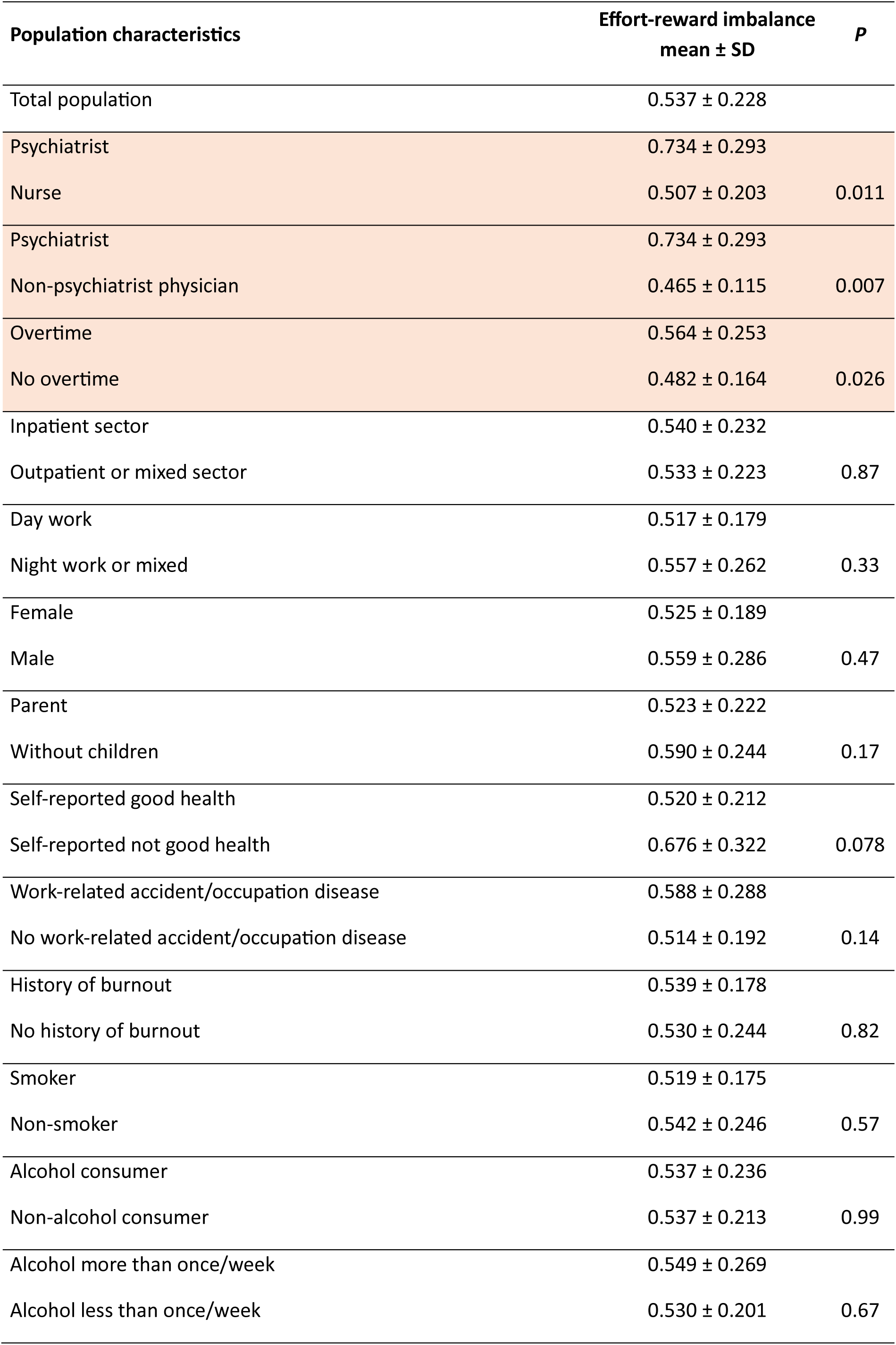
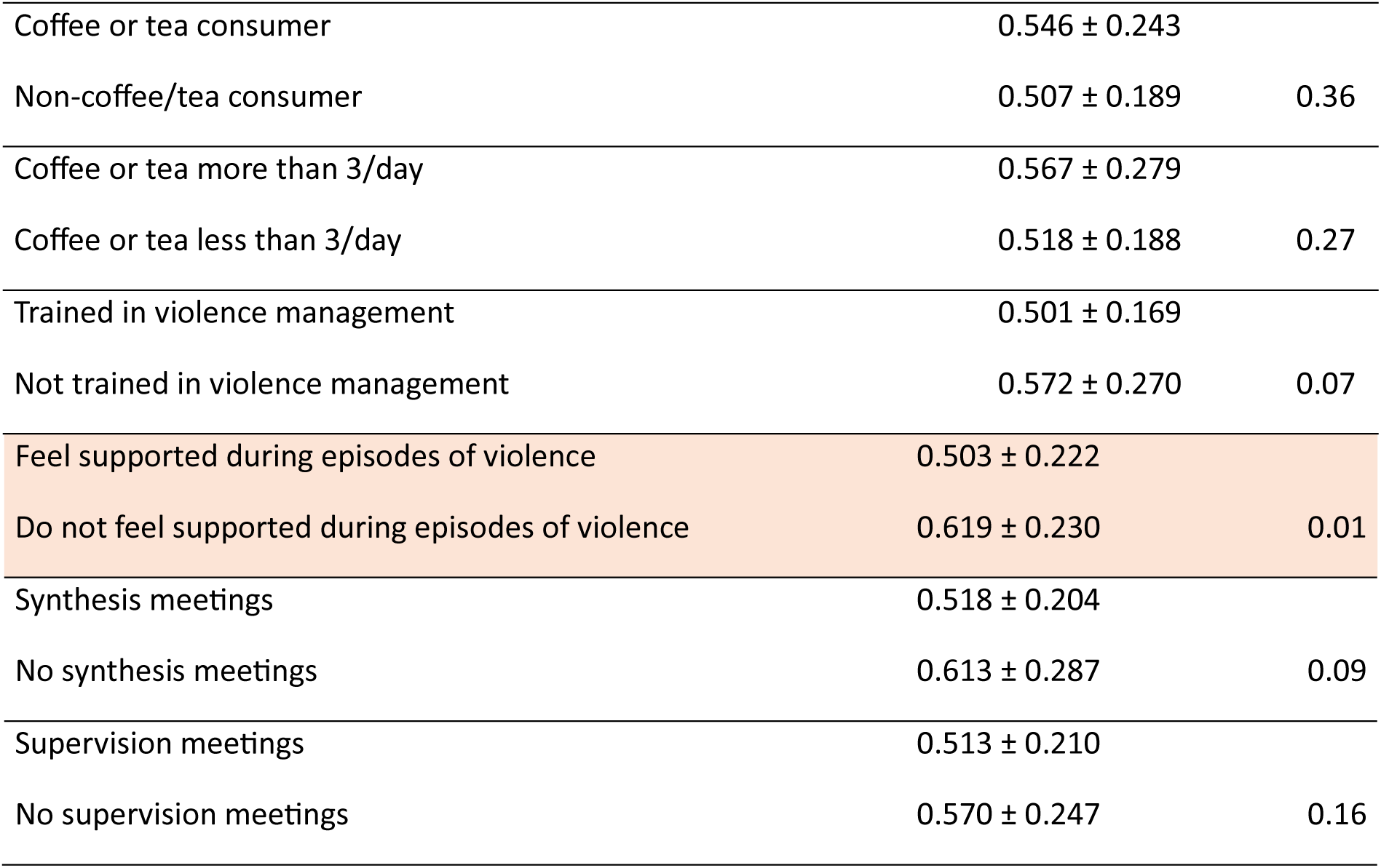
Effort-reward imbalance ratio in population characteristics subgroups.

The ratio was significantly higher for psychiatrists compared to nurses (0.734 ± 0.293 *vs* 0.507 ± 0.203, P=0.011) and non-psychiatrist physicians (0.734 ± 0.293 *vs* 0.465 ± 0.115, P=0.007), as shown in Figure 3.

**Figure 3:**
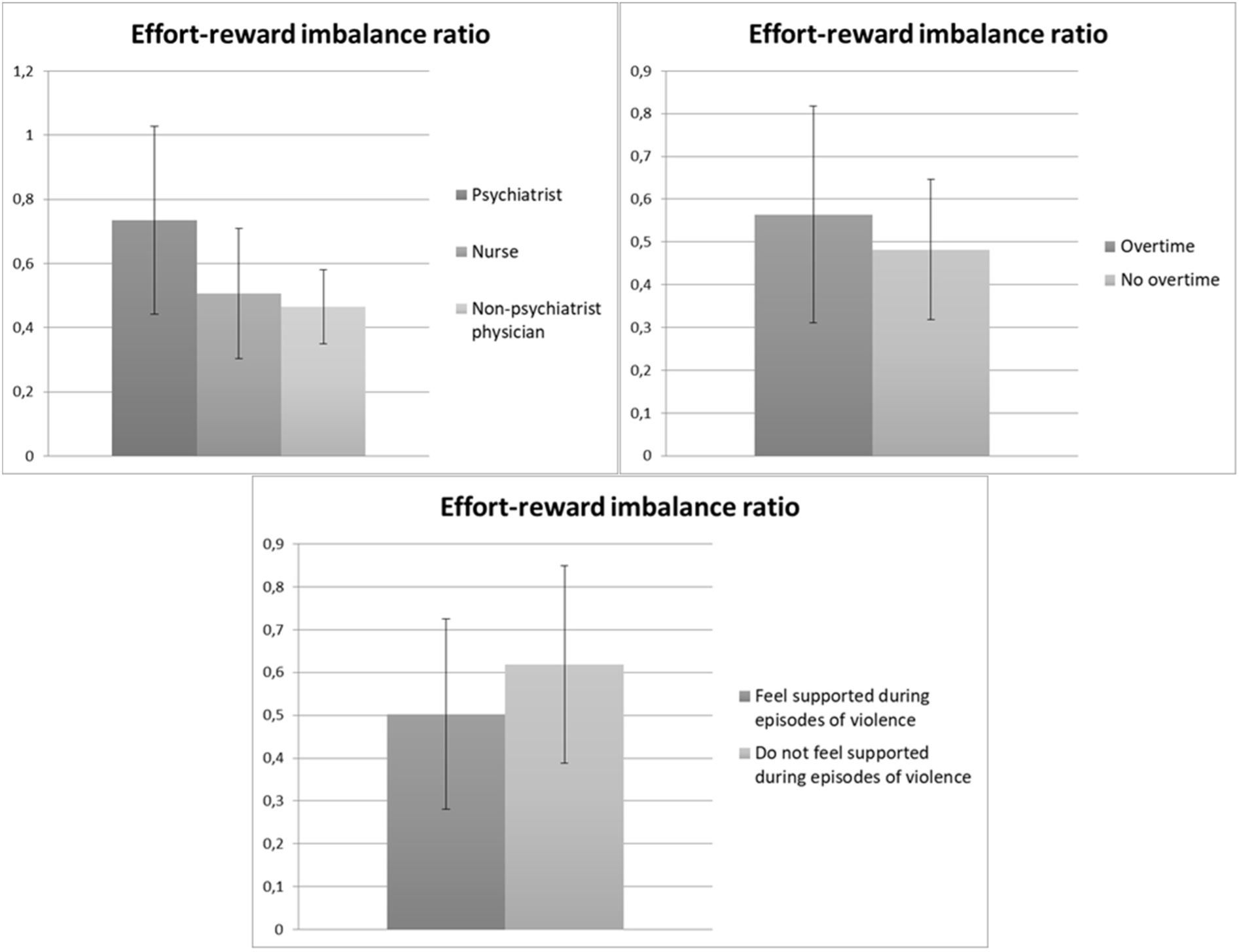
Effort-reward imbalance ratio within the subgroups: profession (top left). overtime (top right) and feeling of support during episodes of violence (bottom).

As compared to the rest of the population, the ratio was also significantly higher in those reporting doing overtime (0.564 ± 0.253 *vs* 0.482 ± 0.164, P=0.026) and lower in those who felt helped during violence episodes (0.503 ± 0.222 *vs* 0.619 ± 0.230, P=0.01).

The analysis of quantitative variables, including age, seniority, and body-mass index did not show any significant correlation.

The analysis of overinvestment and social support scores in population characteristics subgroups is presented in Table 5.

**Table 5:**
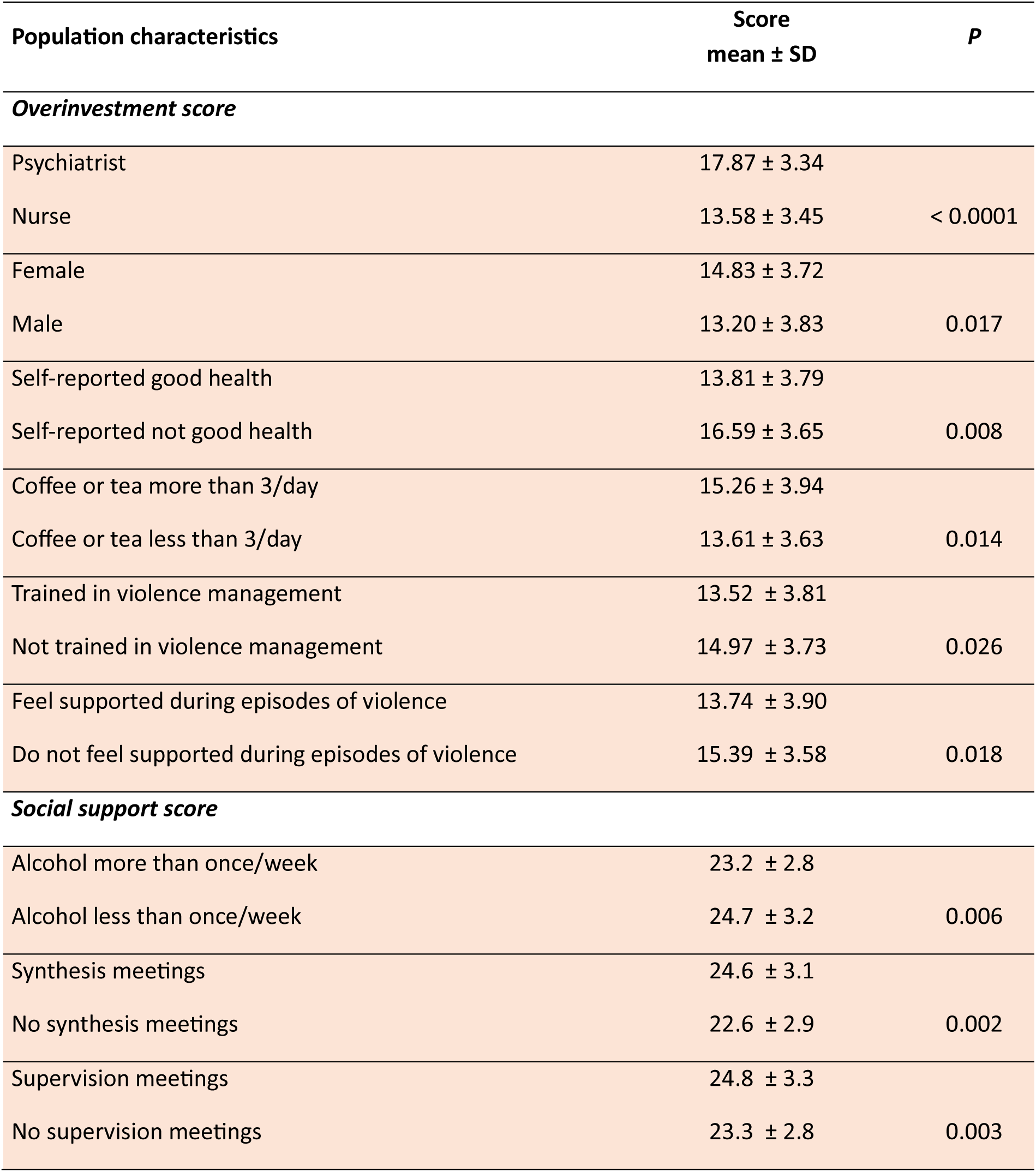
Analysis of overinvestment and social support scores in population characteristics subgroups.

The overinvestment score was significantly higher among psychiatrists as compared to nurses (17.87 ± 3.34 *vs* 13.58 ± 3.45, P<0.0001, among women (14.83 ± 3.72 *vs* 13.20 ± 3.83, P=0.017), and among those not considering themselves in good health (16.59 ± 3.65 *vs* 13.81 ± 3.79, P=0.008).

Consuming coffee or tea more than three times per day was associated with a higher overinvestment score (15.26 ± 3.94 *vs* 13.61 ± 3.63, P=0.014).

This score was significantly higher among those who did not feel sufficiently trained in managing violence (14.97 ± 3.73 *vs* 13.52 ± 3.81, P=0.026) or did not feel helped during violence episodes (15.39 ± 3.58 *vs* 13.74 ± 3.90, P=0.018). These results are illustrated in Figure 4.

**Figure 4:**
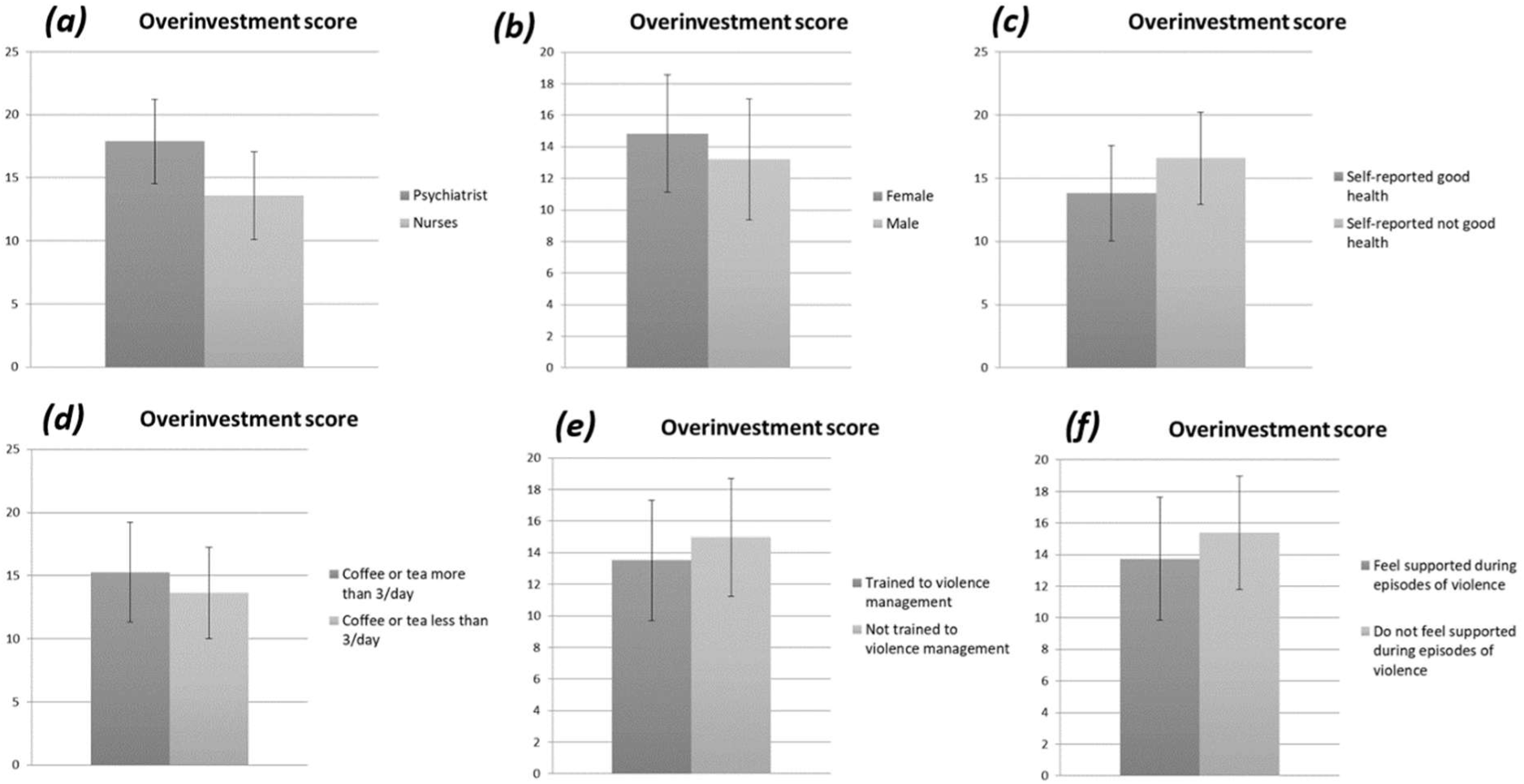
Overinvestment score within the subgroups: (a) profession. (b) gender. (c) self-reported health. (d) coffee or tea consumption. (e) training in violence management and (f) feeling supported during episodes of violence.

The social support score was significantly lower in those who reported consuming alcohol more than once per week (24.7 ± 3.2 *vs* 23.2 ± 2.8, P=0.006).

This score was significantly higher in those who reported participating in synthesis meetings (24.6 ± 3.1 *vs* 22.6 ± 2.9, P=0.002) or supervision meetings (24.8 ± 3.3 *vs* 23.3 ± 2.8, P=0.003). These results are summarized in Figure 5.

**Figure 5:**
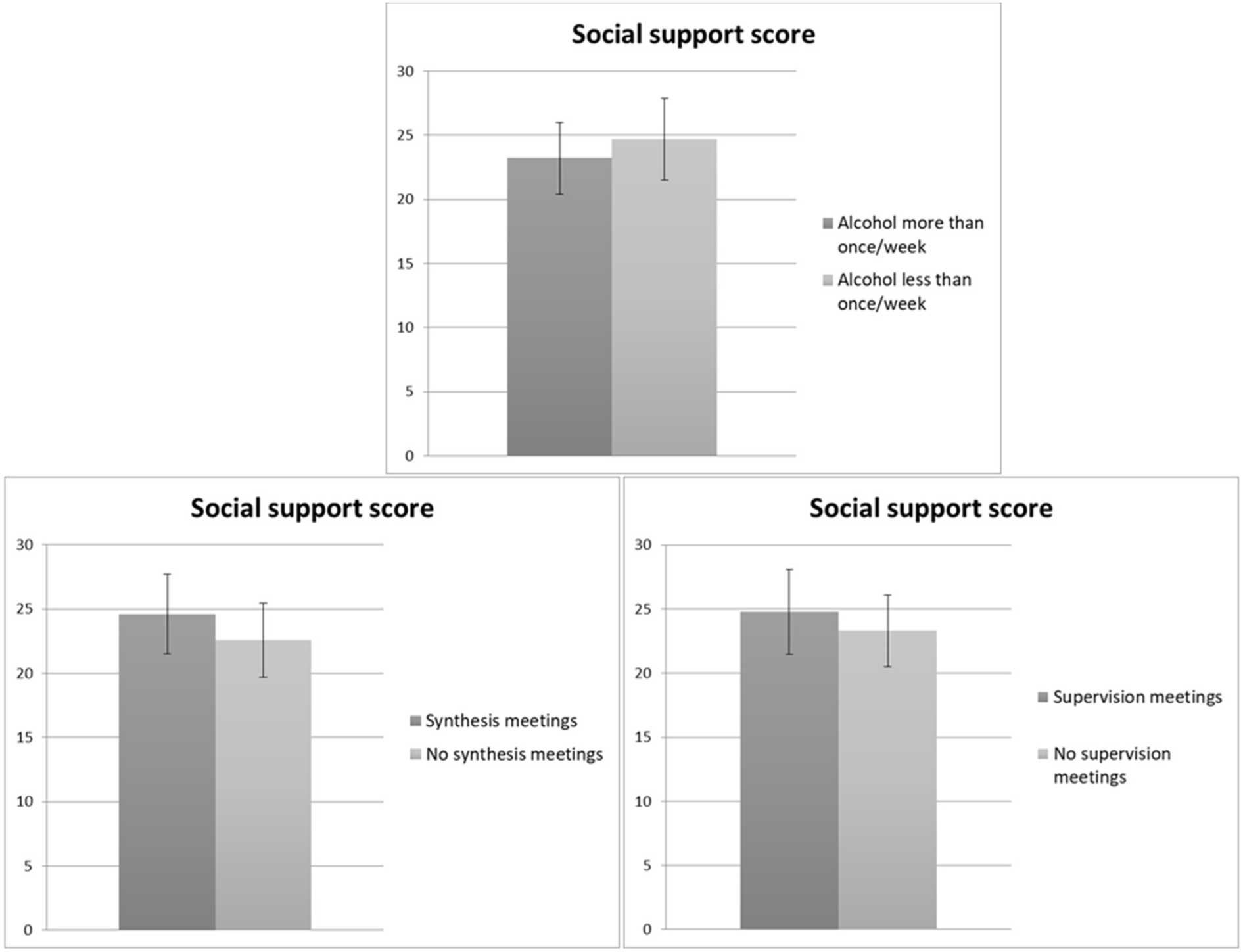
Social support score within subgroups: alcohol consumption (top). synthesis meetings (bottom left). supervision meetings (bottom right).

## Discussion

### Strengths and limitations

The small sample size in our study limits the statistical power of the results, and the single-center design reduces its representativeness. The retrospective nature of the study also lowers the level of evidence. The statistical power in this study was not sufficient to allow for multivariate analysis that could account for confounding factors. For the assessment of psychosocial risks at work, two validated questionnaires were used; however, other questionnaires also exist (21,22). As we used self-administered questionnaires, the responses depend on the respondent’s ability to understand and interpret the questions. This study did not target nursing assistants, who may also be concerned. However, in French psychiatric units, care is mainly provided by nurses.

A strength of this study is that it was conducted across all psychiatric care units in a public French hospital (Table 1).

In our study, the response rate was 65.1%. In comparable studies using self-administered questionnaires with healthcare staff, response rates were 11% (23), 54% (24), and 52.5% (25). A study conducted in Taiwan reported a 98% response rate, but this represented only 36% of the target population (26). A large number of international and French studies did not report response rates (11,16,27–29). This relatively high response rate, which is representative of the target population, may be in relation with the multiple reminders and the in-person presentation of the questionnaire by the investigating physician.

The responding population is representative of the source population in terms of gender distribution (65% women) and average age (45.8 ± 11.2 years), which is comparable to similar studies addressing healthcare workers (23,27). There was no significant difference (P(χ²)=0.82) in the response rates between nurses (63.6%), physicians (70.0%), and supervisors (77.8%).

### Description of the population

Among the respondents, 62% work overtime and 56% work night shifts. To our knowledge, there are no publications on these data in psychiatry. However, a French national study reported that healthcare professionals work 2 hours more than the average French worker, with work constraints felt as “forced” by 70% of them and “disruptive” by 56% (30).

The smoking rate in the study population is 21.7%, which is lower than the rate found in 2012 among psychiatric nurses (31), but similar to the rate among healthcare professionals (20%) in 2022 (30). In our study, 36% of respondents report drinking alcohol more than once a week. A national survey in 2022 found that 41% of healthcare workers and 45% of French people aged 25 to 64 reported such level of consumption (30).

Among the respondents, 88% consider themselves in good health. In a national report, this proportion was 82% for healthcare professionals in 2018, but dropped to 76% in 2022 after the COVID pandemic. For the general French population, this proportion is 85% (87% among working people) (30). A French survey in 2023 showed a trend toward improvement, with 80% of healthcare workers reporting being in good health, but this rate is still lower than the general population (32). Another report in 2023 found the same trend, with 22% of healthcare workers considering their health to be poor or very poor (27). This proportion, slightly better in this study compared to other healthcare workers in France, could be explained by a multidisciplinary team approach that helps managing stress and values professional skills. In the targeted units, a report by the General Controller of Places of Deprivation of Liberty (33) noted that the staff is supportive and well-trained, that there is strong coordination between inpatient and outpatient services for patient care and therapeutic projects, and that dynamic exchanges occur between all categories of staff. The report also emphasized the ethical reflection around professional practices and the integration of activities into patient care. This organization likely contributes to a better work experience and strengthens feeling of good health.

However, the high proportion of healthcare workers considering themselves in good health (88%) contrasts with the high prevalence of “job strain” (37%). This could indicate an underestimation of work-related unhappiness. The Karasek questionnaire may be more sensitive than healthcare workers’ self-perception of their health to detect work-related unhappiness.

In the studied population, 23% of staff regularly take psychotropic medication, which seems higher than the national estimate, ranging from 6 to 13% among healthcare professionals and 8% among French people aged 25 to 64 (27,30).

### Job strain

The thresholds defined in the national Sumer survey from 2003 (34), which surveyed 24,486 employees, are used to compare the results of this study.

The prevalence of “job strain” at 37.4% is higher than that observed in the Sumer survey among male employees (20%) and female employees (28%) with variations ranging from 10% to 36% depending on profession and social category (35). Compared to Sumer, the higher prevalence in our study could be interpreted as an upward trend in exposure to job strain over the past 20 years, or as a particularly high exposure to job strain in the studied population.

In our study, the risk of “job strain” was higher among staff working in inpatient psychiatric settings compared to those in outpatient or mixed settings. This may be linked to the higher intensity of work in inpatient settings, with less stable patients and more demanding work schedules due to the need for continuous care. In the outpatient sector, the consistency of care and reintegration of long-term patients by healthcare workers gives more sense to the work. Current mental health policies organizing constrained outpatient care (36) may contribute to an increased risk of work-related stress in outpatient settings.

Day work appeared to be a protective factor, confirming that night work is more likely to be associated with “job strain”.

Seniority was a protective factor, while age did not significantly affect the occurrence of job strain. This could be due to the healthy worker effect or indicate that cumulated professional experience helps better managing job strain.

The protective role of good health self-perception observed in this study is consistent with the well-established link between job strain and health issues.

A history of work-related accidents or occupational diseases was not associated with the occurrence of job strain. It would be interesting to differentiate between somatic and psychological health troubles, work-related accidents and occupational diseases, and to collect the dates of occurrence for a more precise analysis.

In the studied population, burnout and a history of psychiatric disorder were not associated with job strain. This could be due to appropriate redeployment, the healthy worker effect, or the particularly supportive environment in psychiatric services for those who have experienced psychological distress.

Smoking status or body-mass index were not associated with job strain, which may be related to a cultural shift in stress management, particularly with the use of smartphones.

### Effort-Reward Imbalance

The effort-reward imbalance ratio of 0.537 ± 0.228 is similar to that found in other epidemiological studies conducted in France among hospital staff (37). A European comparison study of nurses found an average ratio of 0.62 ± 0.23 in France in 2011 (15). The proportion of respondents with a risk ratio above 1 is low (4.5% in our sample), similar to other comparable studies (15,37).

Effort-reward imbalance is more pronounced among psychiatrists and among healthcare workers reporting overtime, although these groups were not associated with job strain. Thus, the issue does not seem to lie in the workload but rather in the lack of recognition felt by staff.

This imbalance was also higher among respondents who did not feel adequately supported during episodes of violence. In the literature, barriers to obtaining help in this hospital context are primarily related to tensions with management, with staff sometimes feeling ignored or prevented from speaking up (28). A qualitative analysis of interviews with hospital physicians concluded that improving the organization of care and controlling working hours could enhance physicians’ professional activities (38). No significant association was found with other variables such as age or seniority.

### Overinvestment and social support scores

The overinvestment score was higher among psychiatrists, those consuming coffee or tea more than three times a day, women, and those who did not perceive themselves as being in good health. Training and assistance regarding episodes of violence appear to protect against overinvestment. In 2014, a survey suggested that the professional activities of psychiatrists could be improved by organizing work time to foster patient listening, interprofessional exchanges, and the training of paramedical staff (19).

The social support score was relatively homogeneous in our population. It was lower among healthcare workers who reported consuming alcohol more than once a week, which raises questions about potential causality or consequence. The implementation of dedicated time, such as synthesis meetings and supervision time, was associated with a higher social support score.

## Conclusion

This study found a high prevalence (37%) of “job strain” among psychiatric healthcare workers, with the main risk factors being inpatient work and night work. In contrast, higher seniority and good health self-perception appear to be protective factors. Our results show a high effort-reward imbalance among psychiatrists and healthcare workers who work overtime. These two subgroups report a greater sense of lack of recognition compared to the rest of the study population. In order to reduce the high prevalence of job strain among healthcare workers in psychiatry, national occupational health policies should promote preventive measures addressing the specific risk factors associated with professional practice.

The authors have no conflicts of interest to disclose.

## Data Availability

All data produced in the present work are contained in the manuscript

